# Clonal hematopoiesis of indeterminate potential contributes to accelerated chronic kidney disease progression

**DOI:** 10.1101/2024.06.19.24309181

**Authors:** Caitlyn Vlasschaert, Yang Pan, Jianchun Chen, Elvis Akwo, Varun Rao, James E. Hixson, Michael Chong, Md Mesbah Uddin, Zhi Yu, Mengdi Jiang, Fenfen Peng, Shirong Cao, Yinqiu Wang, Do-Kyun Kim, Adriana M. Hung, Jing He, Manjula Kurella Tamura, Debbie L. Cohen, Jiang He, Changwei Li, Zeenat Bhat, Panduranga Rao, Dawei Xie, Alexander G. Bick, Bryan Kestenbaum, Guillaume Paré, Michael J. Rauh, Adeera Levin, Pradeep Natarajan, James P. Lash, Ming-Zhi Zhang, Raymond C. Harris, Cassianne Robinson-Cohen, Matthew B. Lanktree, Tanika N. Kelly, the CRIC Study Investigators

## Abstract

**Background:** Clonal hematopoiesis of indeterminate potential (CHIP) is a common inflammatory condition of aging that causes myriad end-organ damage. We have recently shown associations for CHIP with acute kidney injury and with kidney function decline in the general population, with stronger associations for CHIP driven by mutations in genes other than *DNMT3A* (non-*DNMT3A* CHIP). Longitudinal kidney function endpoints in individuals with pre-existing chronic kidney disease (CKD) and CHIP have been examined in two previous studies, which reported conflicting findings and were limited by small sample sizes.

**Methods:** In this study, we examined the prospective associations between CHIP and CKD progression events in four cohorts of CKD patients (total N = 5,772). The primary outcome was a composite of 50% kidney function decline or kidney failure. The slope of eGFR decline was examined as a secondary outcome. Mendelian randomization techniques were then used to investigate potential causal effects of CHIP on eGFR decline. Finally, kidney function was assessed in adenine-fed CKD model mice having received a bone marrow transplant recapitulating*Tet2*-CHIP compared to controls transplanted wild-type bone marrow.

**Results:** Across all cohorts, the average age was 66.4 years, the average baseline eGFR was 42.6 ml/min/1.73m^2^, and 24% had CHIP. Upon meta-analysis, non-*DNMT3A* CHIP was associated with a 59% higher relative risk of incident CKD progression (HR 1.59, 95% CI: 1.02-2.47). This association was more pronounced among individuals with diabetes (HR 1.29, 95% CI: 1.03-1.62) and with baseline eGFR ≥ 30 ml/min/1.73m (HR 1.80, 95% CI: 1.11-2.90). Additionally, the annualized slope of eGFR decline was steeper among non-*DNMT3A* CHIP carriers, relative to non-carriers (β -0.61 ± 0.31 ml/min/1.73m^2^, p = 0.04). Mendelian randomization analyses suggested a causal role for CHIP in eGFR decline among individuals with diabetes. In a dietary adenine mouse model of CKD, *Tet2*-CHIP was associated with lower GFR as well as greater kidney inflammation, tubular injury, and tubulointerstitial fibrosis.

**Conclusion:** Non-*DNMT3A* CHIP is a potentially targetable novel risk factor for CKD progression.

## Introduction

Clonal hematopoiesis of indeterminate potential (CHIP) is characterized by the clonal expansion of blood cells carrying somatic mutations in specific driver genes.^1,2^ An age-related disorder, CHIP is rare in the young but its prevalence increases rapidly in older adults, with at least 10% of individuals aged 65 and older affected.^3,4^ Recent studies have identified a causal role for CHIP in several chronic diseases of aging including atherosclerotic cardiovascular disease^5–8^, heart failure^9–13^, gout^14^, liver fibrosis and cirrhosis^15^, osteoporosis^16^, and chronic obstructive pulmonary disorder (COPD).^17^ CHIP is also recognized as a risk factor for acute kidney injury (AKI) severity and non-recovery.^18^ Mouse model evidence and human genetic studies point to inflammation as the key mediator of the CHIP-associated risk in each of these conditions. A hallmark feature of chronic kidney disease (CKD)^19,20^, chronic inflammation confers higher risks of kidney failure in CKD patients. ^12–14^ CHIP has been associated with incident kidney function decline in the general population^21^, though it is not clear whether the inflammatory burden of CHIP would meaningfully intensify the already-inflamed CKD state and affect clinical outcomes. CHIP and longitudinal kidney function outcomes in CKD patients have been examined in two previous cohort studies^22,23^, and results were conflicting. However these studies were limited by small sample sizes and had important differences in CHIP variant assessment and classification.^24^

In this study, we first examine the prospective associations between CHIP and CKD progression events in four large CKD cohorts, totaling 6,216 individuals: the Chronic Renal Insufficiency Cohort (CRIC), the African American Study of Kidney Disease (AASK), subjects with CKD from the BioVU biorepository, and the Canadian study of prediction of death, dialysis and interim cardiovascular events (CanPREDDICT). We then use Mendelian randomization as an orthogonal method to assess the contribution of CHIP to eGFR decline. Finally, we evaluated the effect of experimental *Tet2-*CHIP on kidney function in a mouse model of CKD.

## Methods

### Cohort descriptions

#### Chronic Renal Insufficiency Cohort (CRIC)

CRIC is an on-going observational cohort study of 5,499 adult nephrology patients with baseline eGFR of 20-70 ml/min/1.73m^2^. The first phase of the CRIC study enrolled 3,939 participants (21–74 years) from 2003 to 2008 at seven U.S. clinical centers, with an additional 1,560 participants enrolled during a second recruitment phase between 2013 and 2018. Details regarding the CRIC study design and inclusion criteria were reported previously.^25^ At annual clinical follow-up visits, eGFR was estimated using the race-free creatinine and cystatin C based CRIC equation.^26^ Kidney failure, defined as receipt of maintenance dialysis or a kidney transplant, was ascertained at annual follow-up visits and 6-month interim telephone interviews. Kidney failure events were confirmed by the dialysis unit or hospital chart review and supplemented by information from the United States Renal Data System. Deaths were ascertained by reports from next of kin, hospital records, and periodic searches in the Social Security Administration’s Death Master File, with confirmation by death certificate. CHIP was measured in a subsample of 2,087 CRIC participants who were 65 years of age or older. After excluding participants who failed sequencing (N=75) or were missing CKD progression estimates (N=208) or covariables (n=236), a total of 1,568 participants were included in the current study.

#### African American Study of Kidney Disease (AASK)

The AASK was a randomized controlled trial (RCT) comparing the effect of hypertensive therapies and blood pressure targets on kidney function decline outcomes in African American adults with hypertension and eGFR 20-65 ml/min/1.73m^2^.^27^ 1094 participants were randomized to an intervention (ramipril, metoprolol or amlodipine) and to a blood pressure target (mean arterial pressure (MAP) <90 or MAP 102-107). The main outcomes assessed in the original study were the change in GFR over time (measured by iothalamate clearance) and a major adverse kidney event (MAKE) outcome (i.e., composite 50% eGFR decline, decrease of eGFR by 25 points, kidney failure, or death).^27^ All individuals were followed for up to 6 years as part of the trial, and then for up to 5 years as part of a cohort phase. 836 of the 1094 enrolled participants provided consent for DNA collection, and 209 had DNA available for CHIP-genotyping as part of this study.

### BioVU

BioVU is a biorepository of DNA collected from clinical encounters at Vanderbilt University Medical Center (VUMC) with ongoing sample collection since 2004. Specimen collected in BioVU are linked to de-identified clinical data in the VUMC electronic medical records system. For the purposes of this study, 2445 adults with CKD – defined by the presence of two outpatient eGFR measurements <60 ml/min per 1.73 m^2^ within a minimum of 90 days and a maximum of 365 days apart prior to DNA sampling and a minimum of 2 years and four longitudinal eGFR measurements – were selected.^28^ Serum creatinine values <0.4 and >20 mg/dl were excluded because they likely represented laboratory or data entry errors.

### Canadian study of prediction of death, dialysis and interim cardiovascular events (CanPREDDICT)

CanPREDDICT is an observational cohort enrolling individuals with baseline eGFR of 15 to 45 ml/min/1.73m^2^ who were followed for up to 5 years.^29^ Individuals on immunosuppression for active glomerulonephritis as well as individuals expected to live less than 1 year by their treating physicians were excluded from the original study. A total of 2402 patients were enrolled in the study, and 1550 had DNA available for this study. Significant kidney events and kidney function were tracked at protocolized study visits every 6-12 months.^29^ Individuals who were included in a published pilot study of CHIP in CKD (N = 85) were excluded from the current analysis.^22^

### CHIP ascertainment

Acquired DNA mutations meeting established criteria for CHIP were identified for participants with available DNA in the CRIC (N = 1568), AASK (N = 209), CanPREDDICT (N = 1550), and BioVU (N = 2445) cohorts. The AASK, BioVU, and CRIC cohorts were sequenced using a TWIST panel at Vanderbilt University Medical Center (VUMC). The CanPREDDICT cohort was sequenced using a similar TWIST panel at McMaster University, with gene coverage masked to match the genes covered on the VUMC panel. The average sequencing depth was 547x across CanPREDDICT samples, 671x for CRIC, 618x for AASK, and 703x for BioVU. Samples with poor sequencing coverage (average sequencing depth ≤30) were removed from the analysis.

Putative somatic variants were identified using Mutect2 in all 3 cohorts and filtered using a variant allele fraction (VAF) threshold ≥ 2%, a total sequencing depth of 20 and minimum allele depth of 3. A second somatic variant calling pipeline, DRAGEN variant caller, was used to verify calls in CanPREDDICT since this was a new sequencing panel and several low VAF variants were noted after running Mutect2; only variants identified by both algorithms were kept in the analysis for this cohort. CHIP calls were adjudicated by the lead author and verified by co-authors using previously established standard criteria.^4^ CHIP subtypes were defined by the size of the CHIP largest clone (VAF ≥ 10% and <10%) and by the identity of the mutated gene (i.e., *DNMT3A*-CHIP, non-*DNMT3A* CHIP, *TET2*-CHIP, *ASXL1*-CHIP, and *JAK2*-CHIP).

### Incident outcome ascertainment

The primary outcome was a CKD progression endpoint (composite of 50% eGFR decline or kidney failure, defined as initiation of dialysis or kidney transplantation). Secondary outcomes included CKD progression or death (composite) and slope of eGFR. These outcomes were assessed on an individual level from the date that DNA was obtained (baseline) to end of study follow-up.

### Mendelian randomization

We investigated the effect of genetically-instrumented CHIP on eGFR decline in CKD patients using two-sample Mendelian randomization analyses. Summary statistics used to derive the CHIP instrument were from a large discovery GWAS of 628,388 individuals from the UK Biobank and Geisinger MyCode Community Health Initiative (GHS).^30^ Genetic instruments were independent GWAS-significant SNPs (p<5x10^-8^) for any CHIP that were clumped at an r^2^ of less than 0.1. Summary data for the association of the genetic instruments with eGFR decline were extracted from a meta-analysis of GWAS of eGFR decline among 116,870 participants with CKD (defined by two outpatient eGFR measurements of < 60 ml/min/1.73m^2^) from the Million Veteran Program and BioVU^28^, wherein eGFR decline was defined using the annualized relative slope in outpatient eGFR.

### Mouse studies

#### Animals

All mice were on the C57/Bl6 background. Hematopoietic *Tet2* deficiency was obtained as we have previously reported.^18^ Briefly, recipient mice were lethally irradiated with 9 Gy using a cesium γ source, and bone marrow cells harvested from syngeneic donor femurs and tibias (5 × 10^6^ BM cells in 0.2 ml medium) were administered via retroorbital injection. Mice received either 100% wild type bone marrow (Cd45.2 isotype, WT) or 80% wild type and 20% bone marrow from mice on a C57Bl/6 Cd45.1 with global *Tet2* deletion mice. Mice were studied when the mutant hematopoietic cells were found to have expanded to ∼60% of the total cells, which was determined by flow cytometry. To induce chronic tubulointerstitial injury, mice were fed adenine (containing 0.25% of adenine) in their food for 14 days.^31^ Male mice were used for these studies because they are known to develop more tubulointerstitial fibrosis in response to injury.^32^

#### Glomerular filtration rate (GFR) measurement

FITC-sinistrin (90 mg/kg body weight) was injected retro-orbitally, and the cutaneous fluorescence signal was measured continuously for 1h. The half-life (t_1/2_) of FITC-sinistrin (in minutes) was determined with MPD Studio Software. GFR was adjusted for measured body weight at each time point.

#### Immunoblotting analysis

Whole kidney tissue was homogenized with lysis buffer containing 10 mmol/l Tris–HCl (pH 7.4), 50 mmol/l NaCl, 2 mmol/l EGTA, 2 mmol/l EDTA, 0.5% Nonidet P-40, 0.1% SDS, 100 μ /l Na3VO4, 100 mmol/l NaF, 0.5% sodium deoxycholate, 10 mmol/l sodium pyrophosphate, 1 mmol/l PMSF, 10 μ aprotinin, and 10 μ l leupeptin and centrifuged at 15, 000 x g for 20 min at 4°C. The BCA protein assay kit (Thermo Scientific) was used to measure the protein concentration. Immunoblotting analyses were performed to quantify specific proteins of interest using antibodies to KIM-1 (R&D Systems Cat #AF1817), NGAL (R&D Systems Cat # AF1857), PDGFR-β (R&D Systems Cat # AF1042), α SMA (Abcam Cat #ab21027), fibronectin (Sigma Cat # F3648), and β actin (Cell Signaling Cat # 4967). Immunoblotting was quantitated with Image J software.

#### Quantitative PCR

Total RNAs from kidneys or cells were isolated using Trizol® reagent (Invitrogen). SuperScript IV First-Strand Synthesis System kit (Invitrogen) was used to synthesize cDNA from equal amounts of total RNA from each sample. Quantitative RT-PCR was performed using TaqMan real-time PCR (7900HT, Applied Biosystems). The Master Mix and all gene probes were also from Applied Biosystems. The probes used in the experiments included mouse *Tnf* (Mm99999068), *Il1b* (Mm00434228), *Il6* (Mm00446190), *Ccl2* (Mm00441242), *Col1a1* (Mm00801666), *Col3a1* (Mm01254476), *Acta2* (Mm01546133), *Fn* (Mm01256744), *Havcr1* (KIM-1 Mm00506686)*, Lcn2l* (NGAL Mm01324470)*, Vim* (Mm01333430)*, Cd68* (Mm03047343), F4/80 (Mm Mm00802529) and *Gapdh* (Mm99999915) was used as a normalizer. Realtime PCR data were analyzed using the 2-ΔΔCT method to determine the fold difference in expression.

**Picro-Sirius red stain** was performed according to the protocol provided by the manufacturer (Sigma, St. Louis, MO, USA).

#### Kidney tubular injury score

Periodic acid-Schiff (PAS)–stained slides were used to evaluate tubular injury score. Tubular injury was defined as tubular dilation, tubular atrophy, tubular cast formation, sloughing of tubular epithelial cells or loss of the brush border and thickening of the tubular basement membrane using the following scoring system: Score 0: no tubular injury; Score 1: <10% of tubules injured; Score 2: 10–25% of tubules injured; Score 3: 25–50% of tubules injured; Score 4: 50–74% of tubules injured; Score 5: >75% of tubules injured.

### Statistical analyses

Incident event analyses were assessed using Cox proportional hazards regression models. The secondary outcome of eGFR decline was assessed using a mixed linear regression model, where the repeatedly measured continuous outcome is included as the dependent variable while CHIP, follow-up time, and a CHIP by follow-up time interaction term are included as independent variables. The interaction term and its P-value was used to evaluate the influence of CHIP on eGFR decline over time. In this analysis, individuals are necessarily censored when they develop kidney failure as accurate eGFR measurement is no longer possible. As such, individuals with better kidney function over time contribute more data points to the mixed model regression, with a tendency to skew the results toward a more positive eGFR slope the longer the observation period, as shown in our pilot analysis.^22^ Thus, the first three years of follow-up were examined as the study window for this analysis. Subgroup analyses according to CHIP clone size, CHIP driver genes, diabetes history, and baseline eGFR strata were also conducted. All analyses were adjusted for age, age-squared, sex, baseline eGFR, baseline urine albumin-to-creatinine ratio (or protein-to-creatinine ratio), baseline cardiovascular disease, diabetes, and hypertension, and self-reported race/ethnicity. CRIC, BioVU, and AASK analyses were additionally adjusted for smoking history, which was not available for CanPREDDICT participants. Results were examined for reproducibility across cohorts, and random effects meta-analyses were employed to combine effects across studies.

For Mendelian randomization, the primary analysis was conducted using the conventional multiplicative random-effects inverse variance weighted (IVW) estimator and stratified by diabetes status. As in our previous work, we excluded variants in the telomerase reverse transcriptase (*TERT*) gene as these are known to be associated with several comorbidities and are potentially pleiotropic.^18^ Outliers were investigated using standard approaches including Cook’s distance and residual-versus-leverage plots. Sensitivity analyses were conducted using MR-RAPS and MR-Egger methods. The MR-Egger intercept was used to test for directional pleiotropy. The SNP heritability for the independent SNPs used as genetic proxies for CHIP explained 0.55% of the variance in CHIP after excluding the SNPs in the *TERT* gene.

For animal experiments, statistical analyses were performed with GraphPad Prism 9 (GraphpadSoftware® Inc., La Jolla, CA, US). Data are presented as the mean ± S.E.M. Data were analyzed using 2 tailed Student’s t test or two-way ANOVA followed by Tukey’s or Bonferroni’s post hoc tests. A P value below 0.05 was considered significant.

### Study approval

Research ethics approval for cohort sample sequencing and analyses of the data were obtained from the Vanderbilt University Medical Center (BioVU and AASK cohorts), University of Illinois Chicago (CRIC cohort) and McMaster University (CanPREDDICT cohort) institutional review boards. The Vanderbilt University Medical Centre animal care committee approved all animal study procedures.

## Results

### Baseline characteristics

CHIP was ascertained in a total of 5,772 participants with baseline CKD stages G3-4 across four cohorts using targeted sequencing assays. The cohorts included CRIC (mean baseline eGFR: 46.4 ± 15.5; 85% stage G3), the BioVU biorepository (mean baseline eGFR: 48.0 ± 18.9; 82% stage G3), AASK (mean baseline eGFR: 48.4 ± 12.4; 91% stage G3), and CanPREDDICT (mean baseline eGFR ± SD: 29.3 ± 9.8 ml/min/1.73m^2^; 43% stage G3). Our cohorts represent a racially/ethnically diverse population of patients with CKD, including 1206 African Americans (20.9%) based on self-report. Baseline characteristics are presented in **Table 1**.

**Table 1.** Baseline characteristics for the observational cohorts.

|  | CRIC | BioVU | AASK | CanPREDDICT |
| --- | --- | --- | --- | --- |
| *N* | 1568 | 2445 | 209 | 1550 |
| *CHIP, % (N)* | 27.6 (433) | 18.9 (463) | 27.3 (57) | 26.5 (410) |
| *Non-DNMT3A CHIP, % (N)* | 13.0 (203) | 10.3 (228) | 11.5 (24) | 16.32 (253) |
| *Age, mean ± SD* | 70.2 ± 3.7 | 63.2 ± 13.5 | 60.5 ± 5.9 | 67.5 ± 12.8 |
| *Female, % (N)* | 40.4 (634) | 52.6 (1285) | 35.4 (74) | 37.6 (582) |
| *Race-ethnic group, % (N)* |  |  |  |  |
| *Black* | 40.4 (634) | 13.9 (340) | 100 (209) | 1.5 (23) |
| *White* | 49.7 (779) | 86.1 (2105) | 0 | 90.3 (1400) |
| *Hispanic* | 6.7 (105) | 0 | 0 | 0.3 (5) |
| *Other* | 3.2 (50) | 0 | 0 | 7.9 (122) |
| *Current smoker, % (N)* | 8.4 (132) | 9.4 (199) | 59 (124) | N/A |
| *BMI, mean ± SD* | 31.8 ± 6.6 | N/A | 30.6 ± 6.4 | N/A |
| *Estimated GFR, mean ± SD* | 46.4 ± 15.5 | 48.0 ± 18.9 | 48.4 ± 12.4 | 29.3 ± 9.8 |
| *Urinary PCR (mg/mg), mean ± SD* | 0.57 ± 1.41 | N/A | 0.31 ± 0.53 | 1.02 ± 1.81 |
| *Urinary ACR, mean ± SD* | 449.4 ± 910.1 | 355.9 ± 517.3 | N/A | 774.1 ± 1480.0 |
| *Hypertension, % (N)* | 92.4 (1449) | 235.7 (658) | 100 (209) | 97.5 (1511) |
| *Use of ACEi/ARBs, % (N)* | 71.2 (1116) | 87.4 (2137) | 34.4 (72) | 73.4 (1138) |
| *Diabetes, % (N)* | 54.1 (848) | 29.5 (722) | 0 | 47.3 (733) |
| *CVD, % (N)* | 42.7 (670) | 26.4 (645) | 51.7 (108) | 32.3 (501) |

CHIP variants were identified in 433 of 1568 individuals (27.6%) in CRIC, 463 of 2445 individuals (18.9%) in BioVU, 57 of 209 individuals (27.3%) in AASK, and 410 of 1550 individuals (26.5%) in CanPREDDICT. Approximately 50% of identified variants were in the CHIP driver gene *DNMT3A*, and variants in genes other than *DNMT3A* (i.e., non-*DNMT3A* CHIP) comprised the other half. The mean and median clone sizes (quantified as variant allele fractions, or VAF) for the largest clone per person were: 8.8% and 4.7% per person in CRIC, 8.3% and 4.5% in BioVU, 6.0% and 3.5% in AASK, and 10.4% and 5.1% in CanPREDDICT (Supplemental Figure 1). Approximately half of all detected CHIP variants were small clones (VAF ≤ 5%), reflecting the sensitivity of the targeted sequencing approach.

### CHIP and incident kidney disease progression

Our primary endpoint was a composite of 50% eGFR decline or kidney failure (*i.e.,* initiation of dialysis or receipt of kidney transplant). When all CHIP gene mutations were included in the meta-analysis, CHIP was not associated with the primary endpoint (HR 1.17, 95% confidence interval (CI): 0.94–1.46; **Figure 1a** and **Supplemental Table 1**). However, when CHIP gene subtypes were considered separately, non-*DNMT3A* CHIP was associated with significantly higher risk of the primary composite endpoint (HR 1.59, 95% CI: 1.02-2.47) while *DNMT3A*-CHIP was not (HR 0.98, 95% CI: 0.80-1.19). These results were similar in sensitivity analyses considering only incident kidney failure as an endpoint (**Supplementary Table 2**).

**Figure 1.**
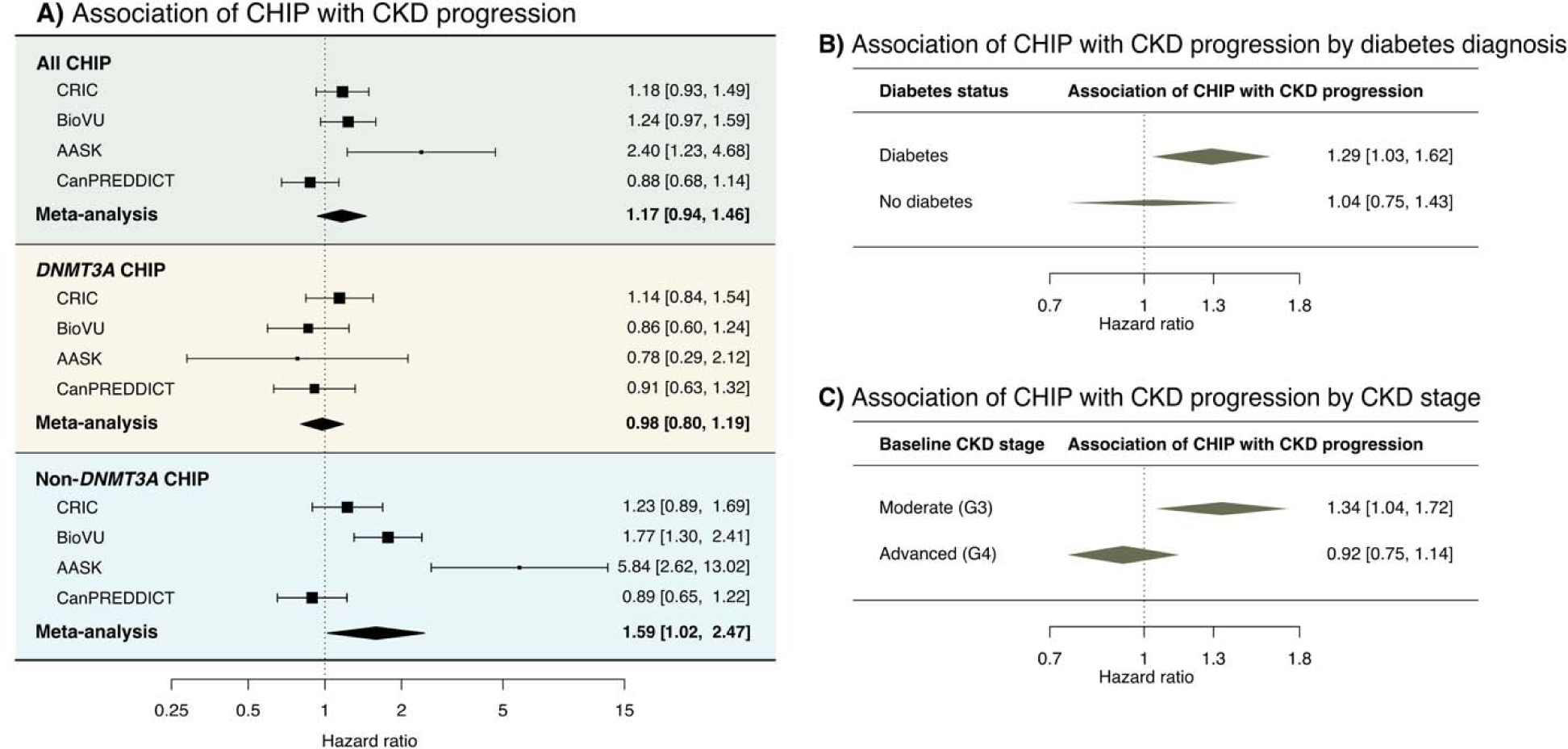
**A)** Non-*DNMT3A* CHIP is associated with CKD progression defined as a composite of incident 50% eGFR decline or kidney failure. **B)** An association between CHIP and CKD progression is observed in individuals with baseline moderate (stage G3) but not advanced (stage G4) CKD. **C)** There is a significant interaction between CHIP and baseline diabetes status with respect to incident CKD progression.

Since CHIP is associated with death, and premature death is a common adverse event for individuals with CKD, we examined the composite risk of incident kidney disease progression or death in our cohorts as a sensitivity analysis. CHIP was associated with a higher risk of this outcome (HR 1.14, 95% CI: 1.00-1.29), and the risk was greater for non-*DNMT3A* CHIP (HR 1.53, 95% CI: 1.12-2.08) and among individuals with diabetes (HR 1.33, 95% CI: 1.12-1.57). The full results for the composite kidney disease progression or death endpoint are reported in **Supplementary Table 3**.

In subgroup analyses, CHIP was associated with CKD progression in individuals with baseline diabetes (HR 1.29, 95% CI: 1.03-1.62) but not those without diabetes (HR 1.04, 95% CI: 0.75-1.43; **Figure 1b**). The subgroup interaction term for diabetes was not significant, though there was marked heterogeneity among the non-diabetic cohort subsets (p = 0.03) driven by inclusion of the AASK cohort (**Supplemental Figure 2**). The AASK cohort was the smallest cohort (∼10% the size of other cohorts) and had unique features compared to the other cohorts: it was a clinical trial as opposed to an observational cohort, and only African Americans without diabetes were enrolled. In a sensitivity analysis excluding the AASK cohort, the interaction term for diabetes was significant (p = 0.03).

Across baseline eGFR strata, the effect sizes were more pronounced for individuals with baseline CKD stage G3 (HR 1.34, 95% CI: 1.04–1.72 for all CHIP and HR 1.80, 95% CI: 1.11–2.90 for non-*DNMT3A* CHIP; **Figure 1c**) and indistinguishable from the null in individuals with a more advanced baseline CKD stage G4 (HR 0.92, 95% CI: 0.75-1.14 for all CHIP and HR 1.04, 95% CI: 0.76–1.42 for non-*DNMT3A* CHIP).

### CHIP and rate of eGFR decline

We then assessed whether CHIP was associated with a faster rate of eGFR decline using mixed model regressions of serial eGFR measurements. In a random effects meta-analysis, CHIP was not associated with the slope of eGFR decline (β -0.17 ± 0.19 ml/min/1.73 m^2^ per year, p = 0.36). Non-*DNMT3A* CHIP was also not associated with eGFR slope (β -0.42 ± 0.37 ml/min/1.73 m^2^ per year, p = 0.25), though there was significant heterogeneity between the cohorts (p = 0.04; **Figure 2a**), with a negative slope for all cohorts except CanPREDDICT (the cohort with the lowest average baseline eGFR; **Table 1**). In an analysis examining only individuals with baseline eGFR ≥ 30 ml/min/1.73 m^2^, non-*DNMT3A* CHIP was associated with significantly steeper slope of eGFR decline (β -0.61 ± 0.30 ml/min/1.73 m^2^ per year, p = 0.04; **Figure 2b**). The full results for the eGFR decline analyses are reported in **Supplemental Table 4**.

**Figure 2.**
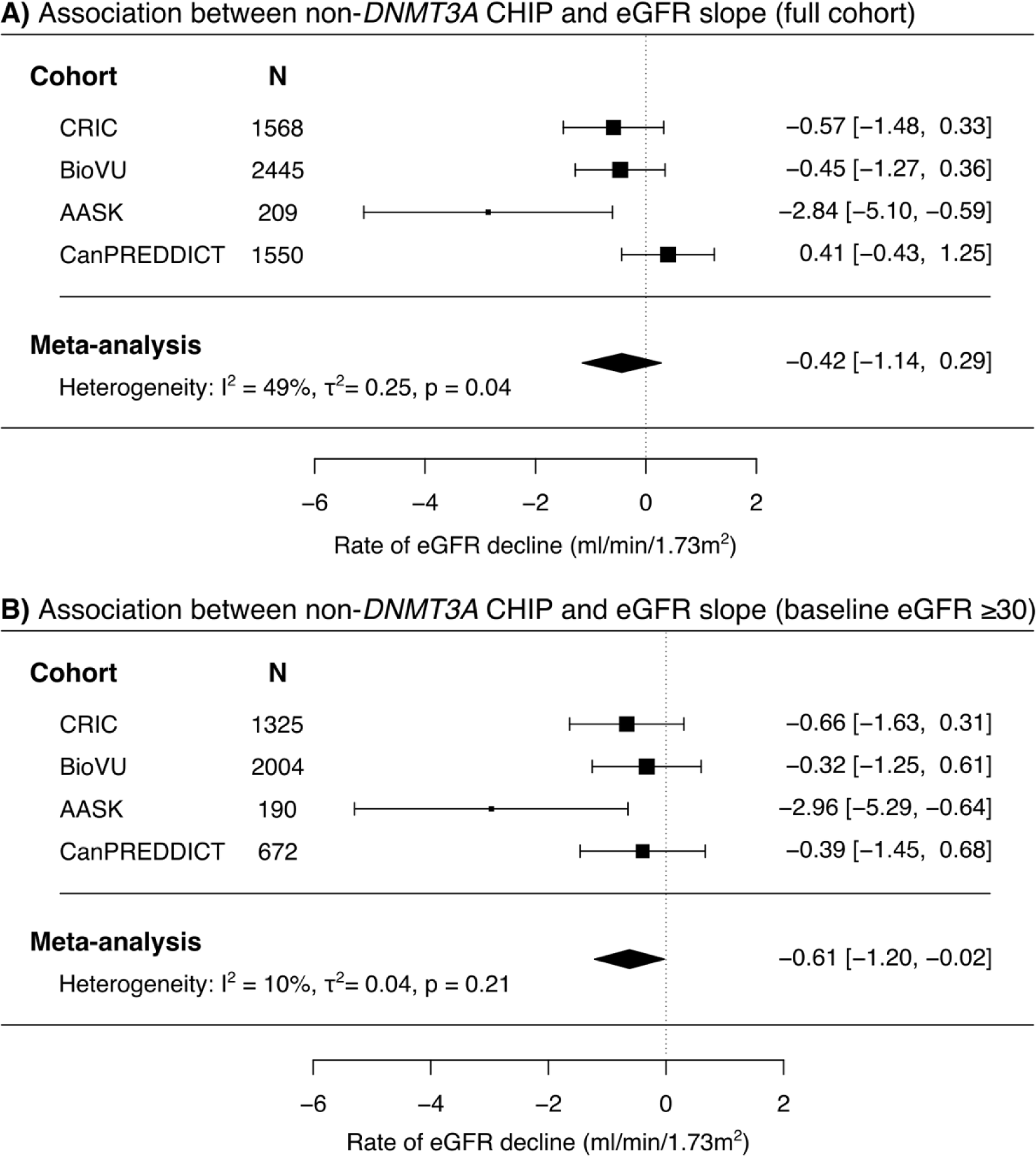
Non-*DNMT3A* CHIP is associated with a steeper slope of eGFR decline among individuals with baseline eGFR ≥ 30 ml/min/1.73m^2^ (73% of all participants).

### Mendelian randomization

As shown in Table 2, a genetic instrument for CHIP was significantly associated with a 0.25% per-year faster eGFR decline (p=0.04) in diabetic CKD patients while the findings were mostly null for patients with nondiabetic kidney disease (p-interaction = 0.05; **Table 2**). The MR estimate in diabetic patients was consistent across other MR methods including MR-RAPS (β - 0.23, 95% CI: -0.49–0.03) and MR-Egger (β -0.36, 95% CI: -1.1–0.35) albeit with wider 95% confidence intervals (**Supplemental Figure 3**). The p-value for the MR-Egger intercept test was 0.77 suggesting no evidence of directional pleiotropy.

**Table 2.** Mendelian randomization estimates for the effect of the genetic instrument for CHIP on annualized relative eGFR slope in CKD patients stratified by diabetes status.

|  | Mendelian Randomization Estimate |  |  |  |
| --- | --- | --- | --- | --- |
|  | Random-Effects IVW<sup>1</sup> |  | MR-RAPS<sup>2</sup> |  |
|  | Beta<sup>3</sup> (95% CI) | *P* | Beta<sup>3</sup> (95% CI) | *P* |
| Diabetes | -0.25 (-0.50, -0.01) | 0.04 | -0.23 (-0.49, 0.03) | 0.08 |
| No Diabetes | 0.01 (-0.11, 0.13) | 0.84 | 0.02 (-0.09, 0.13) | 0.71 |
| P-interaction | 0.05 |  | 0.09 |  |

<sup>1</sup>Multiplicative random-effects inverse-variance-weighted estimate
<sup>2</sup>Mendelian randomization using robust adjusted profile score
<sup>3</sup>Beta represents the percent change in the eGFR per year per fold-increase in the odds of CHIP (all subtypes).

### CHIP and kidney disease severity in a mouse model of chronic kidney disease

We assessed the effect of CHIP on CKD severity in a mouse model where high-dose dietary adenine is used to induce CKD.^31^ As in previously described methods^18^, we created a mouse model of *TET2*-CHIP by performing a bone marrow transplant of 20% CD45.2^+^ *Tet2*^-/-^ cells and 80% CD45.1^+^ *Tet2*^+/+^ cells in irradiated mice as well as paired controls that received a transplant of 100% CD45.1^+^ *Tet2*^+/+^ cells. These mice are hereto forth referred to as *Tet2^-/-^* and WT mice, respectively. After two weeks of dietary adenine administration, *Tet2^-/-^* mice had significantly lower glomerular filtration rates compared to WT mice (57.2 ± 10.2 vs. 98.4 ± 13.1 µl/min, **Figure 3a**). Quantitative PCR indicated increased kidney mRNA expression of macrophage markers (*F4/80*, *Cd68*) and inflammatory cytokine markers (*Il1b, Il6, Tnf, Ccl2*) in adenine-treated *Tet2^-/-^* mice compared to adenine-treated WT mice (**Figure 3b**), and quantitation of F4/80 and Gr-1 immunostaining indicated increased macrophages and neutrophils in the kidneys of *Tet2^-/-^* mice. (**Figure 3c**). *Tet2^-/-^* mice also had increased mRNA for the tubule injury markers (*Kim-1, Ngal;* **Figure 3d**) and an increased tubular injury score (**Figure 3e**). *Tet2^-/-^* mice expressed increased markers of fibrosis (Acta2, *Col1a1, Col3a1, Vim, and Fn;* **Figure 3f**). Picrosirius red staining (**Figure 3g**) and α-SMA immunostaining (**Suppl. Figure 4**) confirmed significantly increased areas of fibrosis in *Tet2^-/-^* kidneys compared to WT kidneys. Immunoblotting confirmed that *Tet2^-/-^* kidneys expressed significantly increased KIM-1, NGAL, a-SMA and fibronectin, as well as increased PDGFR-β compared to WT kidneys (**Figure 3h)**

**Figure 3.**
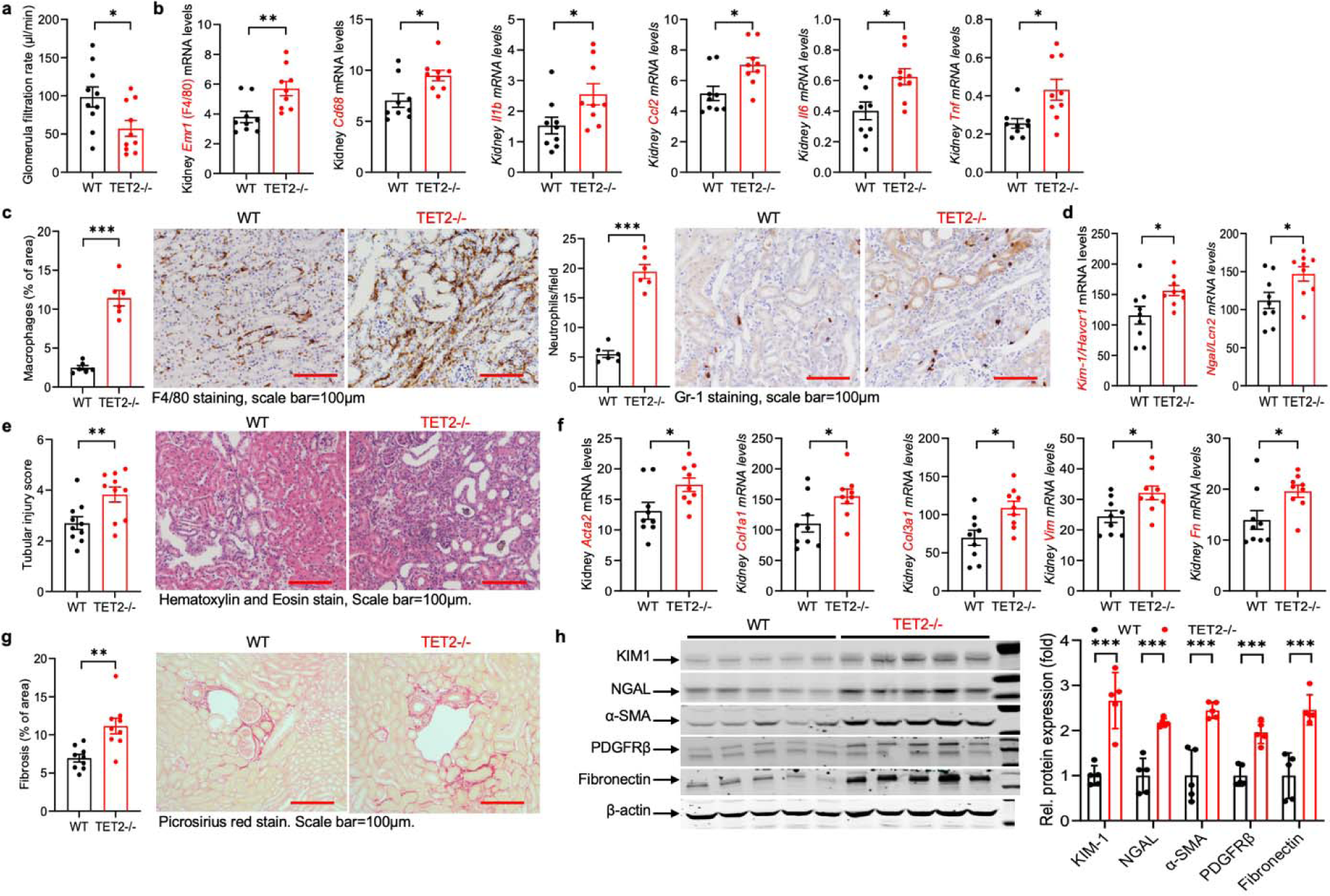
The response of mice with hematopoietic deletion of *Tet2* to adenine-induced chronic kidney injury. **A)** Evaluation of glomerular filtration rate (GFR) at day 14 on adenine diet in *Tet2*^−/−^ and WT mice (*n* = 10 mice each). **B)** Kidney mRNA for macrophage markers *Emr1*/F4/80 and *Cd68* mRNA as well as the proinflammatory cytokines *Il1b*, *Ccl2*, *Il6* and *Tnf* 14 days after adenine diet in *Tet2*^−/−^ and WT mice (*n* = 9 mice each). **C)** Representative images and quantification of F4/80 macrophage and Gr-1 neutrophil immunohistochemistry (IHC) 14 days after adenine diet in *Tet2*^−/−^ and WT mice (*n* = 6 mice each). **D)** Kidney mRNA for *Ngal1/Lcn2* and *Kim-1/Havcr1* 14 days after adenine diet in *Tet2*^−/−^ and WT mice (*n* = 6 mice each). **E)** Tubular injury score 14 days after adenine diet in *Tet2*^−/−^ and WT mice (*n* = 10 mice each). **F)** Kidney mRNA of profibrotic markers *Acta2*, *Col1a*, *Col3a1*, *Vim* and *Fn* 14 days after adenine diet in *Tet2*^−/−^ and WT mice (*n* = 9 mice each). **G)** Representative images of kidney histology stained with Picrosirius red and quantification of kidney interstitial collagen 14 days after adenine diet in *Tet2*^−/−^ and WT mice (*n* = 9 mice each). **H)** Quantification of kidney KIM1, NGAL, α-SMA, PDGFRβ and Fibronectin protein expression in the kidneys of *Tet2*^−/−^ and WT mice 14 days after adenine diet (*n* = 5 mice each). Data were analyzed using a two-tailed Student’s *t*-test and are presented as the mean ± s.e.m. \**P* < 0.05, \*\**P* < 0.01, \*\*\**P* < 0.001.

## Discussion

CHIP is defined by the presence of an expanded clonal white blood cell population caused by acquired mutations in myeloid cancer driver genes. Driver gene identity and size of the clonal population are key determinants of the risk of progression to overt hematologic malignancy and end-organ damage, with non-*DNMT3A* CHIP and larger clones being given more weight in malignancy risk prediction tools.^33^ In the present work, we identify that non-*DNMT3A* CHIP is associated with a greater risk of kidney function decline in individuals with CKD, both when examining incident 50% eGFR decline or kidney failure events and annualized eGFR slopes. In Mendelian randomization analysis, a genetic predisposition for CHIP development was associated with a faster eGFR decline in those with CKD and diabetes. Additionally, in a *Tet2*-CHIP mouse model – the most common type of non-*DNMT3A* CHIP – dietary adenine administration led to more pronounced kidney functional impairment, inflammatory cell infiltration into kidney parenchyma, increased cytokine expression, and development of renal fibrosis when compared to mice without CHIP mutations fed the same diet. These findings are in line with our pilot study^22^ and with our work identifying non-*DNMT3A* CHIP as a risk factor for incident kidney function decline in the general population as well as for impaired recovery after AKI.^18,21^

In subgroup analyses, we find differences in associations by baseline diabetes status, and in Mendelian randomization analyses, we find a greater effect of CHIP on eGFR decline among those with diabetes. These results indicate that the kidney implications of CHIP may be potentiated in the setting of diabetes. CHIP – particularly non-*DNMT3A* CHIP subtypes – has been linked with incident type 2 diabetes diagnoses in humans and with greater insulin resistance in mouse models.^34,35^ In addition to its direct effects on the kidney via pro-inflammatory macrophage parenchymal infiltration^18^, given the link between CHIP and diabetes, one could hypothesize that CHIP may worsen diabetic glucose control and exacerbate microvascular disease. We also find that effect of CHIP is larger among individuals with moderate CKD (baseline eGFR 30-60 ml/min/1.73 m^2^) compared to those with advanced CKD (baseline eGFR ≤ 30 ml/min/1.73 m). The effect of traditional risk factors for CKD including hypertension and diabetes are dampened in those with advanced CKD, which suggests that chronic conditions including CHIP may have less of a discernable effect on progression to kidney failure in more advanced CKD.

A strength of our meta-analysis is that all participants have CKD meeting guideline definitions (based on multiple eGFR measurements), and the 5,772 total participants include a range of CKD etiologies and severities. We selected *a priori* the primary and secondary outcomes that we felt would meaningfully capture CKD progression across this wide range of baseline eGFRs, informed by our pilot study^22^, and we include several sensitivity analyses in the Supplementary Tables. Limitations of our study include that we had limited power to examine all subgroups of interest, including etiologies other than diabetes and driver gene subgroups other than *DNMT3A*-*versus* non-*DNMT3A* CHIP. Notably, the lack of causal effect in Mendelian randomization analyses for individuals without diabetes may be reflective of a lack of statistical power to detect an effect owing to the lower rate of CKD progression among non-diabetics (as exemplified in Table S1 of our paper). Additionally, our targeted sequencing panel does not capture other types of clonal hematopoiesis, including mosaic loss of the Y chromosome (mLOY), which appears to have similar health implications in males as CHIP.^36–39^

In our CKD mouse model, *Tet2*-CHIP was associated with heightened inflammatory responses in the kidneys, including greater neutrophil and macrophage kidney infiltration and pro-inflammatory cytokine expression therein, with consequent kidney scarring and loss of function. This inflammatory model of organ damage is similar to that which has been described for CHIP involvement in cardiovascular, liver, and lung diseases.^5,6,15,17^ Inflammatory cytokine blockade is a proposed treatment strategy to mitigate CHIP-related morbidity and mortality. In an exploratory analysis of the CANTOS trial – which evaluated the role of canakinumab (a monoclonal antibody against IL-1β in preventing recurrence of cardiovascular events in individuals with recent myocardial infarction and evidence of ongoing inflammation (high sensitivity C-reactive protein ≥ 2mg/L)^40^ – individuals with *TET2*-CHIP had a greater benefit from canakinumab compared to individuals without CHIP (HR 0.38, 95% CI: 0.15-0.96).^41^ Additionally, individuals with a common genetic variant in the *IL6R* gene that dampens IL-6 signalling across the lifetime (*IL6R* p.Asp358Ala) have been shown to be protected from CHIP-related cardiovascular disease and stroke in multiple studies.^7,8,42,43^ Monoclonal antibodies targeting IL-6 are being evaluated in individuals with advanced CKD for numerous indications, including cardiovascular disease and anemia refractory to erythropoietin therapy^44,45^, and have shown favourable safety profiles thus far. Individuals with CHIP – particularly non-*DNMT3A* CHIP and/or large clones – may especially benefit from pro-inflammatory cytokine blockade.

In conclusion, non-*DNMT3A* CHIP is a risk factor for the progression of CKD, especially diabetic kidney disease. *Tet2*-CHIP mouse models point to macrophage infiltration and dysregulated inflammatory activity as a causal mechanism. As with other organ systems, proinflammatory cytokine blockade may be a viable therapeutic strategy to mitigate CHIP-related kidney injury.

## Data Availability

The CHIP calls for the observational cohorts will be released via Zenodo alongside publication of the paper.

## Disclosure statement

Funding for the CRIC Study was obtained under a cooperative agreement from National Institute of Diabetes and Digestive and Kidney Diseases (NIDDK; grants U01DK060990, U01DK060984, U01DK061022, U01DK061021, U01DK061028, U01DK060980, U01DK060963, U01DK060902 and U24DK060990). Funding for the ancillary CHIP study in CRIC was supported by R01DK125782. In addition, this work was supported in part by the Perelman School of Medicine at the University of Pennsylvania Clinical and Translational Science Award (National Institutes of Health/National Center for Advancing Translational Sciences [NIH/NCATS] UL1TR000003), Johns Hopkins University (UL1 TR-000424), University of Maryland (GCRC M01 RR-16500), Clinical and Translational Science Collaborative of Cleveland (UL1TR000439 from the NCATS component of the NIH and NIH roadmap for Medical Research), Michigan Institute for Clinical and Health Research (MICHR; UL1TR000433), University of Illinois at Chicago (CTSA UL1RR029879), Tulane COBRE for Clinical and Translational Research in Cardiometabolic Diseases (P20 GM109036), Kaiser Permanente (NIH/National Center for Research Resources [NCRR] UCSF-CTSI UL1 RR-024131), Department of Internal Medicine, University of New Mexico School of Medicine (R01DK119199).

A portion of the data reported here have been supplied by the United States Renal Data System (USRDS). The interpretation and reporting of these data are the responsibility of the author(s) and in no way should be seen as an official policy or interpretation of the U.S. government.

The dataset(s) used for the BioVU analyses described were obtained from Vanderbilt University Medical Center’s BioVU which is supported by numerous sources: institutional funding, private agencies, and federal grants. These include the NIH funded Shared Instrumentation Grant S10RR025141, S10OD025092, and S10OD017985; and CTSA grants UL1TR002243, UL1TR000445, and UL1RR024975. Genomic data are also supported by investigator-led projects that include U01HG004798, R01NS032830, RC2GM092618, P50GM115305, U01HG006378, U19HL065962, R01HD074711; and additional funding sources listed at https://victr.vumc.org/biovu-funding/.

This research was also supported by the National Institute of Diabetes and Digestive and Kidney Diseases (Grant R01DK132155 to C.R.C., B.K., R.H. and A.B.).

## Author contributions statement

C.V. co-designed the study, led CHIP variant identification in all cohorts, performed the incident event analyses for the CanPREDDICT and AASK cohorts, performed the meta-analyses, created the figures, and drafted the manuscript. Y.P. co-designed the study, performed the incident event analyses for the CRIC cohort, and edited the manuscript. C.R.C. and E.A. performed the BioVU incident event analyses as well as the Mendelian randomization analyses, which J.H. and A.M.H. assisted with. J.C. led the mouse model experiments, which M.J., F.P., S.C. and Y.W. assisted with and M.-Z.Z., R.C.H. oversaw. V.R., M.C., M.M.U., Z.Y., G.P., A.G.B. and P.N. coordinated the targeted CHIP sequencing for the prospective cohorts as well as bioinformatic processing of CHIP calls. D.-K.K., J.H., M.K.T., D.L.C., J.H., C.L., Z.B., P.R., D.X., and the CRIC Study Investigators designed the CRIC study and provided critical input insofar as the analysis of this cohort. A.L. designed the CanPREDDICT study and provided critical input insofar as the analysis of this cohort. Finally, C.R.C., M.B.L. and T.N.K. co-designed the study, oversaw the described work while J.E.H., B.K., M.R. and J.P.L. were involved in providing critical feedback throughout the process; each of these authors edited the manuscript.

