## Supplemental Figures for "Clonal hematopoiesis of indeterminate potential contributes to accelerated chronic kidney disease progression"

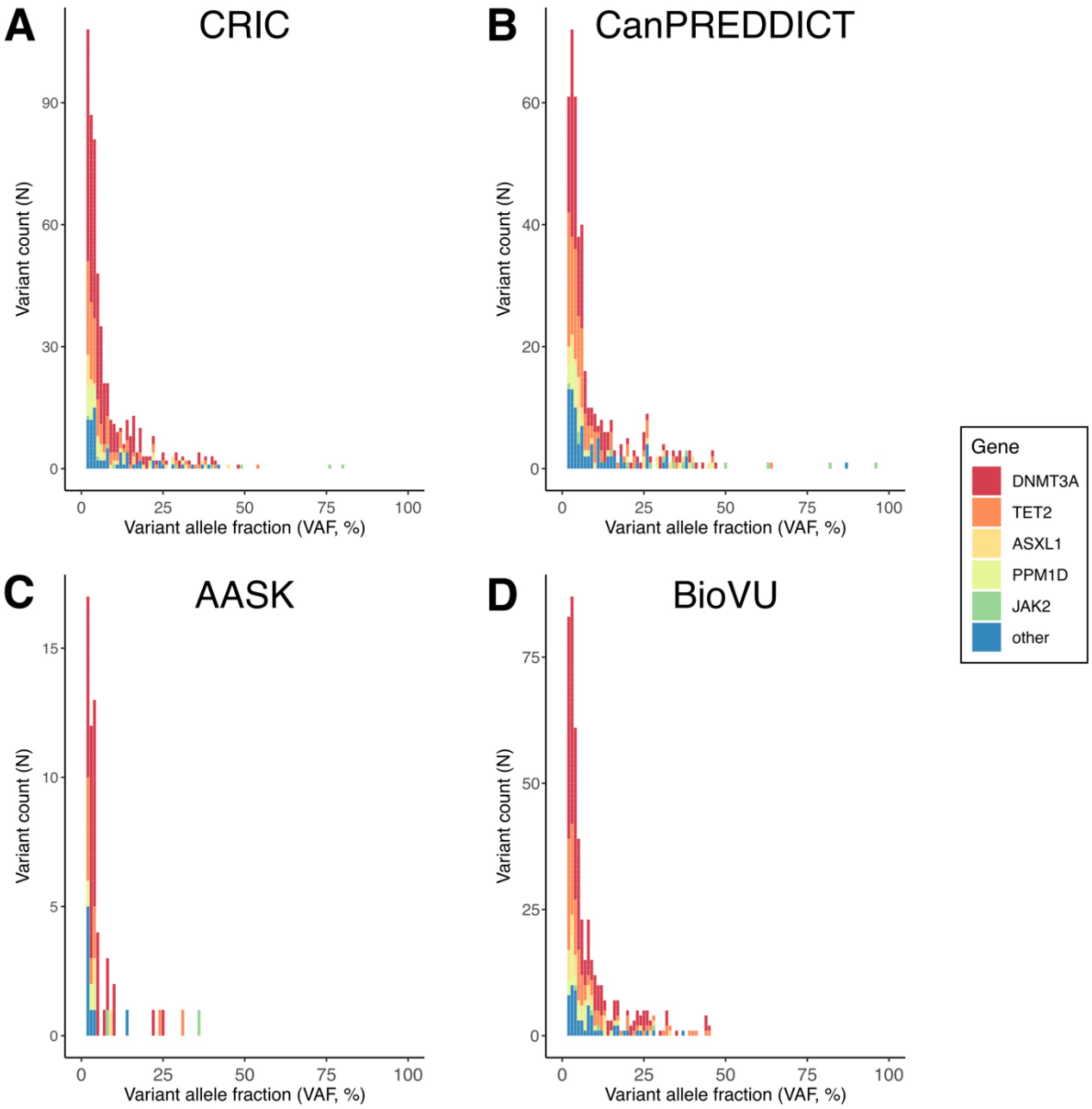


**Supplemental Figure 1.** Distribution of CHIP mutations identified in each cohort by clone size and mutated gene identity.

| Study | TE | seTE | | 95%−CI | Weight (common) | Weight (random) |
|---|---|---|---|---|---|---|
| Subgroup = Diabetes | | | | | | |
| CRIC | 1.34 | 0.1520 | 1.34 | [1.05; 1.64] | 22.5% | 15.5% |
| BioVU | 1.70 | 0.2071 | 1.70 | [1.29; 2.11] | 12.1% | 14.3% |
| CanPREDDICT | 1.00 | 0.1700 | 1.00 | [0.67; 1.33] | 17.9% | 15.2% |
| Common effect model | | | 1.31 | [1.11; 1.50] | 52.5% | -- |
| Random effects model | | | 1.33 | [0.95; 1.72] | -- | 45.0% |
| Heterogeneity: $I^2 = 71\%$, $t^2 = 0.0822$, $p = 0.03$ | | | | | | |
| Subgroup = No diabetes | | | | | | |
| CRIC | 0.94 | 0.2090 | 0.94 | [0.53; 1.35] | 11.9% | 14.3% |
| BioVU | 1.02 | 0.1630 | 1.02 | [0.70; 1.34] | 19.5% | 15.3% |
| AASK | 2.40 | 0.3410 | 2.40 | [1.73; 3.07] | 4.5% | 11.1% |
| CanPREDDICT | 0.71 | 0.2110 | 0.71 | [0.30; 1.12] | 11.7% | 14.2% |
| Common effect model | | | 1.05 | [0.85; 1.26] | 47.5% | -- |
| Random effects model | | | 1.23 | [0.53; 1.92] | -- | 55.0% |
| Heterogeneity: $I^2 = 84\%$, $t^2 = 0.4492$, $p < 0.01$ | | | | | | |
| **Common effect model** | | | **1.19** | **[1.05; 1.33]** | **100.0%** | **--** |
| **Random effects model** | | | **1.26** | **[0.89; 1.64]** | **--** | **100.0%** |

Heterogeneity: $I^2 = 79\%$, $t^2 = 0.2138$, $p < 0.01$
Test for subgroup differences (fixed effect): $c_1^2 = 3.14$, df = 1 ($p = 0.08$)
Test for subgroup differences (random effects): $c_1^2 = 0.07$, df = 1 ($p = 0.79$)

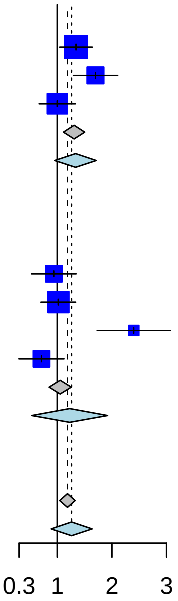


**Supplemental Figure 2.** Risk of CKD progression by diabetes status – subgroup interaction analysis.

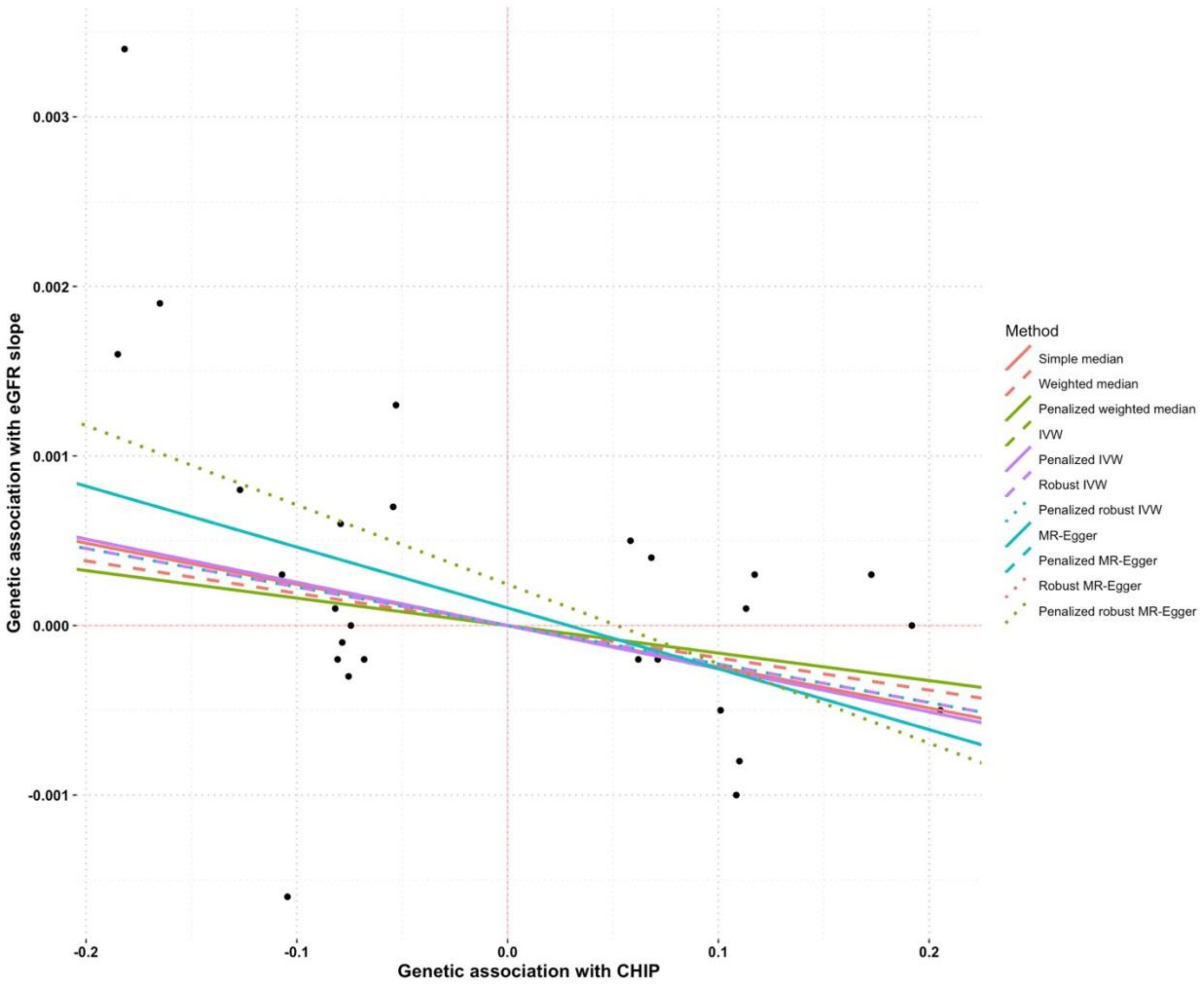


**Supplemental Figure 3.** Estimates of the association between CHIP and eGFR slope among CKD patients with diabetes using different Mendelian randomization techniques.

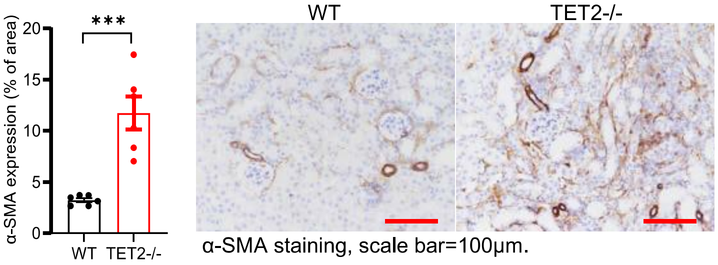


**Supplemental Figure 4.** *Tet2*$^{-/-}$ mice had increased kidney fibrosis compared to WT mice as evidenced by $\alpha$-SMA immunostaining.
